# Age and body mass index affect fit of spirometry GLI references in schoolchildren

**DOI:** 10.1101/2021.10.07.21264678

**Authors:** Rebeca Mozun, Cristina Ardura-Garcia, Eva S. L. Pedersen, Jakob Usemann, Florian Singer, Philipp Latzin, Alexander Moeller, Claudia E. Kuehni, on behalf of the LUIS Study Group

## Abstract

**Background:** References from the Global Lung Function Initiative (GLI) are widely used to interpret children’s spirometry results. We assessed fit for healthy schoolchildren.

**Methods:** LuftiBus in the school (LUIS) is a population-based cross-sectional study done from 2013-2016 in the canton of Zurich, Switzerland. Parents and children aged 6-17 years answered questionnaires about respiratory symptoms and lifestyle. Children underwent spirometry in a mobile lung function lab. We calculated GLI-based z-scores for FEV_1_, FVC, FEV_1_/FVC, and FEF_25-75_ for healthy White participants. We defined appropriate fit to GLI references by mean values ±0.5 z-scores. We assessed if fit varied by age, body mass index, height, and sex using linear regression models.

**Results:** We analysed data from 2036 children with valid FEV_1_ measurements of which 1762 also had valid FVC measurements. The median age was 12.2 years. Fit was appropriate for children aged 6-11 years for all indices. In adolescents aged 12-17 years, fit was appropriate for FEV_1_/FVC (mean: -0.09; SD: 1.02) z-scores, but not for FEV_1_ (mean: -0.62; SD: 0.98), FVC (mean: -0.60; SD: 0.98), and FEF_25-75_ (mean: -0.54; SD: 1.02). FEV_1_, FVC, and FEF_25-75_ z-scores fitted better in children considered overweight (means: -0.25, -0.13, -0.38) than normal weight (means: -0.55, -0.50, -0.55; *p*-trend: <0.001, 0.014, <0.001). FEV_1_, FVC, and FEF_25-75_ z-scores depended on both age and height (*p* interaction: 0.034, 0.019, <0.01).

**Conclusion:** GLI-based FEV_1_, FVC, and FEF_25-75_ z-scores do not fit White Swiss adolescents well. This should be considered when using reference equations for clinical decision making, research and international comparison.

**Take home message:** Our study suggests GLI-based FEV_1_, FVC, and FEF_25-75_ z-scores over detect abnormal lung function in Swiss adolescents, and more so among slimmer adolescents, which has important implications for clinical care, research, and international comparisons.

## Introduction

Standardized global spirometry reference equations are important for interpreting spirometry results, supporting clinical decision-making, and allowing comparison among research findings. To adequately interpret spirometry results and avoid misclassification in clinical care and research, lung function references must fit the local population [1]. The Global Lung Function Initiative (GLI) collected large amounts of spirometry data from healthy non-smoking populations around the world and developed these reference equations [2]. GLI references are now widely used in clinics and studies worldwide. An ideal fit for GLI references with a local population is defined by mean z-scores of spirometry indices equal to 0 with a standard deviation of 1; however, a shift of up to ±0.5 z-scores of the mean is considered acceptable due to sampling variations [3-5]. GLI references account for ages 3-95, ethnicities, height, and sex [2]. Thus, z-scores of spirometry indices should not be affected by these factors. GLI references do not adjust for body composition assessed by weight or body mass index (BMI), though these factors may explain some variability in z-score results in children [6, 7] and excessive weight gain is reported to adversely affect lung growth [8, 9].

Standardizing lung function references during adolescence is particularly complex [10] because physiological changes in lung growth and body structure may occur at different ages for males and females of the same ethnicity [11-13]. European studies showed that GLI references fitted children from the France, Norway, Spain, and the United Kingdom (UK) [14-17], yet only partially fitted children from Italy and Germany [18, 19]. At present, we know little about the factors that contribute to the poor fit for some populations. The fit of GLI references had not been assessed for Swiss children. In this study, we assessed GLI reference equation fit for healthy White schoolchildren in Switzerland and investigated factors that may affect fit.

## Methods

### Study design and setting

LuftiBus in the school (LUIS) is a population-based cross-sectional study of children aged 6-17 years and their respiratory health from 2013-16 in the canton of Zurich, Switzerland (ClinicalTrials.gov: NCT03659838). Zurich is the most populated canton in Switzerland, and it includes many persons born in other countries. The methodology of LUIS has been described [20]. All schools in the canton of Zurich were invited to participate. If the head of a school agreed, trained lung function technicians visited the school in a bus with equipment to measure lung function. Parents completed a detailed questionnaire at home; children answered a questionnaire via interview with technicians at school and underwent lung function testing. The ethics committee of the canton of Zurich approved the study (KEK-ZH-Nr: 2014-0491) and informed consent was obtained from parents and children prior to participation.

### Selection of the study population

We included children with available parent and child questionnaires. To obtain a sample of healthy children, we excluded those with parent-reported wheeze in the past year, use of inhaled corticosteroids (ICS) in the past year or lifetime doctor’s diagnosis of asthma (Figure 1). We excluded children who reported smoking at least once per week and those who reported cough or a cold on the day of the measurement. These are standard criteria to define healthy participants (with some variations between studies) [6, 19, 21, 22]. We excluded children with ethnicities other than White because there were too few to assess fit [1]. We also excluded children with implausible height or body mass index (BMI) z-scores (> 4 or < -4 z-scores), which probably was the result of data recording errors during height and weight measurements. We did quality control checks of lung function measurements, as previously described [20]. We excluded children with invalid lung function measurements, whose flow-volume curves had signs of hesitation at the start of expiration, submaximal effort, early termination of expiration, or cough or glottis closure before the first second of expiration for FEV_1_ and after the first second of expiration for FVC.

**Figure 1:**
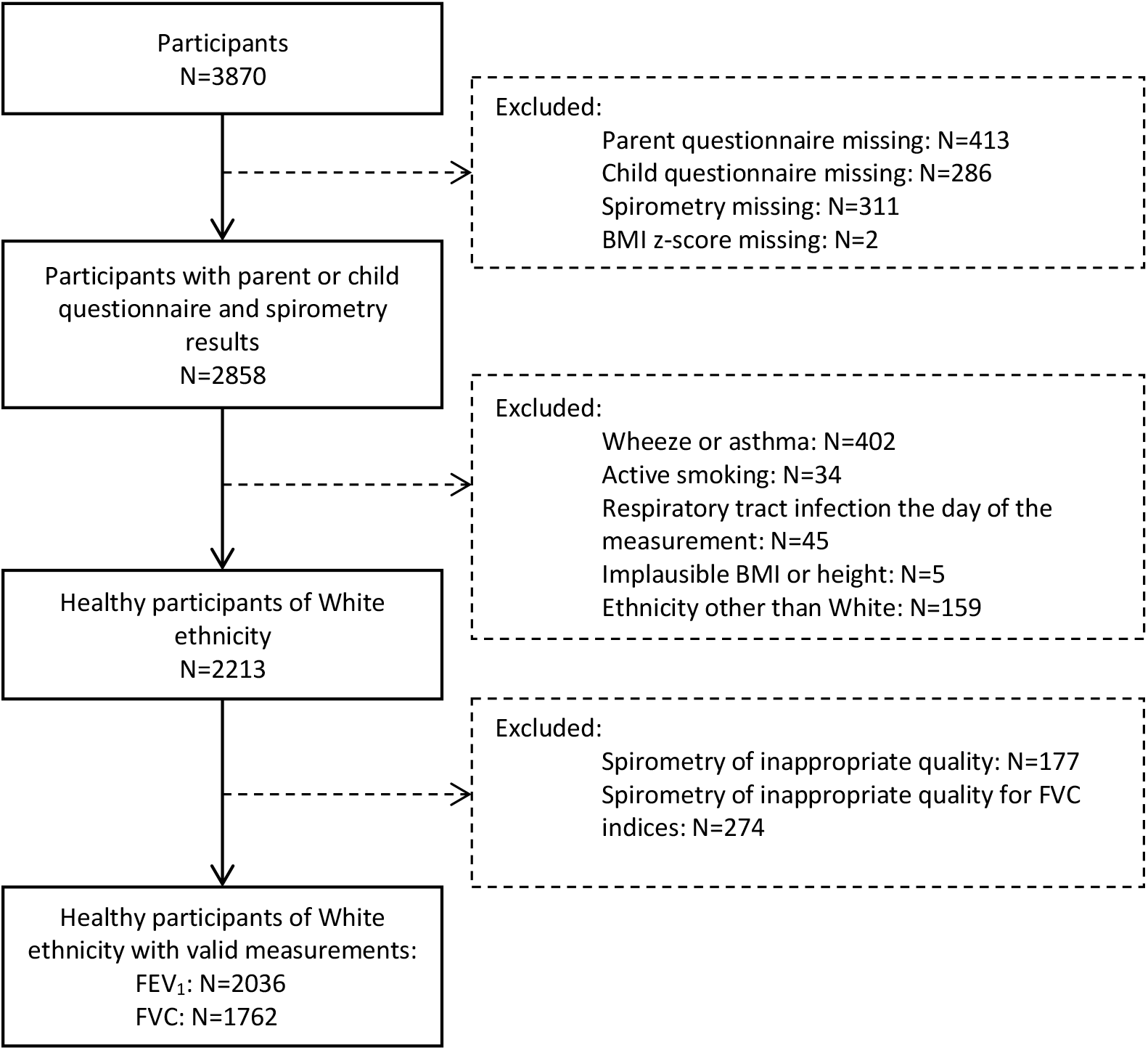
Flow-chart showing the selection of the study population. Wheeze or asthma: parent reported wheeze in the past 12 months; use of inhaled corticosteroids in the past 12 months; or asthma diagnosed by a doctor ever in life. Active smoking: child reported smoking cigarettes, shishas, or e-cigarettes at least once per week. Sick the day of the measurement: child reported strong cough or cold the day of the measurement. Spirometry of inappropriate quality: We also excluded results for FVC-related parameters when we identified signs of early termination of expiration or cough or glottis closure after the first second of expiration in flow-volume curves. Abbreviations: FEV_1_ = forced expiratory volume in 1 second. FVC = forced vital capacity. BMI = body mass index.

### Study procedures

The questionnaire for parents was paper-based and completed at home [20]. It asked about diagnoses, family and household characteristics and lifestyle, medication, and respiratory symptoms. The interview questionnaire for children was shorter and focused on respiratory symptoms, presence of cough or a cold on the day of the measurement, and active smoking. Questions were based on the International Study of Asthma and Allergies in Childhood and the Leicester Respiratory Cohort studies questionnaires [23, 24].

For spirometry we used Masterlab (Jaeger, Würzburg, Germany). Trained technicians conducted spirometry according to ERS/ATS standards [25]. As main outcomes, we used forced expiratory volume in 1 second (FEV_1_), forced vital capacity (FVC), FEV_1_/FVC, and forced expiratory flow between 25 and 75% of the FVC (FEF_25-75_). We applied GLI ethnicity-specific reference equations for Whites to produce z-scores for spirometry indices using the GLI desktop software [26]. Children’s standing height and weight were measured without shoes by the technicians at school according to standard [27]. We calculated BMI z-scores and height-for-age z-scores based on WHO references [27]. We classified BMI z-scores into 4 categories (underweight: <-2 z-scores; normal weight: ≥-2 to <1 z-scores; overweight: ≥1 to <2 z-scores; obese: ≥2 z-scores) [27].

### Statistical analysis

We calculated the mean and 95% confidence interval (CI) for z-scores of spirometry indices. Deviations of the mean outside the range of -0.5 to +0.5 z-scores are generally considered physiologically meaningful [3, 5, 19]. We described the proportion of the population below lower limits of normal ([LLN]; i.e., <-1.645 z-scores) and its 95% CI. Per definition, 5% of the population should fall below the LLN. To address systematic deviations in the fit of GLI references by age, we stratified by 2 age groups: children younger than 12 years and adolescents aged 12 years or older [19]. We chose this age cut-off because puberty begins at different ages for males and females, yet it has usually begun in both sexes by age 12 [28]. To allow further comparisons, the online supplement also shows results stratified by 4 age groups: 6-9, 10-11, 12-13, and 14-17 years [19].

We studied associations of GLI-based z-scores for FEV_1_, FVC, FEV_1_/FVC, and FEF_25-75_ with age, BMI z-scores, height, and sex using scatterplots and univariable linear regression models. We tested for age-height, age-sex, and height-sex interactions using multivariable regression models for each spirometry parameter and compared models with and without interaction terms using likelihood ratio tests.

We performed 3 sensitivity analyses. In the first, we used stricter criteria to define healthy children; in the second, we used looser criteria; and in the third, we excluded children with migration backgrounds (i.e., ethnically White children who neither they nor their parents were born in Switzerland [see online supplement, Table S2]).

We used STATA (version 16, Stata Corp, College Station, TX, USA) for statistical analysis and graphs. We followed STROBE reporting guidelines [29].

## Results

### Study population and sample selection

The LUIS study included 3870 children from 37 schools in the canton of Zurich. From these 3870 children, we analysed data from 2213 children who had parent completed questionnaires and child answered questionnaires, spirometry, and anthropometric measurements, were ethnically White, and healthy (Figure 1). After quality control of flow-volume curves, we included data from 2036 children. All had valid FEV_1_ and 1762 had valid FVC measurements. The median age was 12.2 years (range 6.3-17.0, interquartile range 9.6-14.0) and 49% were males (Table 1). Most children were born in Switzerland (n=1814, 90%) but only two-thirds of their parents were (table 1). Compared to WHO references, our population had a similar BMI distribution (mean BMI z-scores: 0.04) but was slightly taller (mean height-for-age z-scores: 0.56).

**Table 1:** Characteristics of the whole study population and the study sample.

|  | **Whole**<br>**population**<br>**N=2858**<br>**n (%)** | **Sample included**<br>**for analysis**<br>**N=2036**<br>**n (%)** |
| --- | --- | --- |
| Sex, male | 1409 (49%) | 995 (49%) |
| Sex, female | 1449 (51%) | 1041 (51%) |
| Age of the child in years, median (range) | 12.6 (6.0, 17.2) | 12.2 (6.3, 17.0) |
| Age groups |  |  |
| 6–11 years | 1268 (44%) | 975 (48%) |
| 12–17 years | 1590 (56%) | 1061 (52%) |
| Height in cm, median (range) | 156 (105-195) | 154 (105, 194) |
| Height-for-age z-scores, mean (SD) | 0.56 (1.01) | 0.56 (0.98) |
| BMI z-scores, mean (SD) | 0.06 (1.14) | 0.04 (1.10) |
| BMI groups |  |  |
| Underweight (z-scores <-2) | 99 (3%) | 61 (3%) |
| Normal weight (z-scores >-2, <1) | 2168 (76%) | 1573 (77%) |
| Overweight (z-scores >1, <2) | 449 (16%) | 310 (15%) |
| Obesity (z-scores >2) | 142 (5%) | 92 (5%) |
| Child born in Switzerland* | 2522 (88%) | 1815 (90%) |
| Mother originally from Switzerland* | 1833 (64%) | 1338 (67%) |
| Father originally from Switzerland* | 1876 (66%) | 1362 (68%) |
| Swiss SEP, median (range) | 70.2 (28.0, 98.5) | 70.4 (28.0, 98.5) |
| Parent history of asthma* |  |  |
| No | 2404 (84%) | 1772 (87%) |
| Mother | 236 (8%) | 130 (6%) |
| Father | 164 (6%) | 102 (5%) |
| Both | 18 (1%) | 9 (<1%) |
| Wheeze, past 12m* | 230 (8%) | - |
| Use of inhaled corticosteroids, past 12m* | 146 (5%) | - |
| Physician diagnosed asthma ever* | 254 (9%) | - |

Abbreviations: SEP = socioeconomic position index; BMI = body mass index. Height-for-age and BMI z-scores based on WHO references. \*Information obtained from the parent questionnaires.

### Fit of GLI references

For children aged 6–11 years, fit was appropriate for FEV_1_ (mean: -0.37; SD: 0.94), FVC (−0.26; 0.98), and FEF_25-75_ (−0.49; 0.94). For adolescents aged 12–17 years, the fit for all 3 outcomes was poorer: mean FEV_1_ was -0.62 (SD: 0.98), FVC was -0.60 (0.98), and FEF_25-75_ -0.54 (1.02) (Figures 2 and 3, Table S1). Fit of FEV_1_/FVC z-scores was appropriate for both age groups (mean z-scores: 6-11 years: -0.14; SD: 1.06; 12-17 years: -0.09; 1.02). The proportion of participants with mean z-scores below the LLN was above the expected 5% for the 6–11 year age group for FEV_1_ (9%; 95% CI: 7-11), FVC (8%; 6-10), and FEF_25-75_ (11%; 9-13). Among adolescents, this was even more pronounced for FEV_1_ (15%; 13-17), FVC (14%; 12-16), and FEF_25-75_ (14%; 11-16). For FEV_1_/FVC, proportions below LLN were close to 5% for males aged 6–11 years (4%; 3-7) and females (7%; 5-10) and males (7%; 5-10) aged 12-17 years, yet not for females aged 6–11 years (9%; 7-12). Findings were very similar in the 3 sensitivity analyses (Tables S2 and S3).

**Figure 2:**
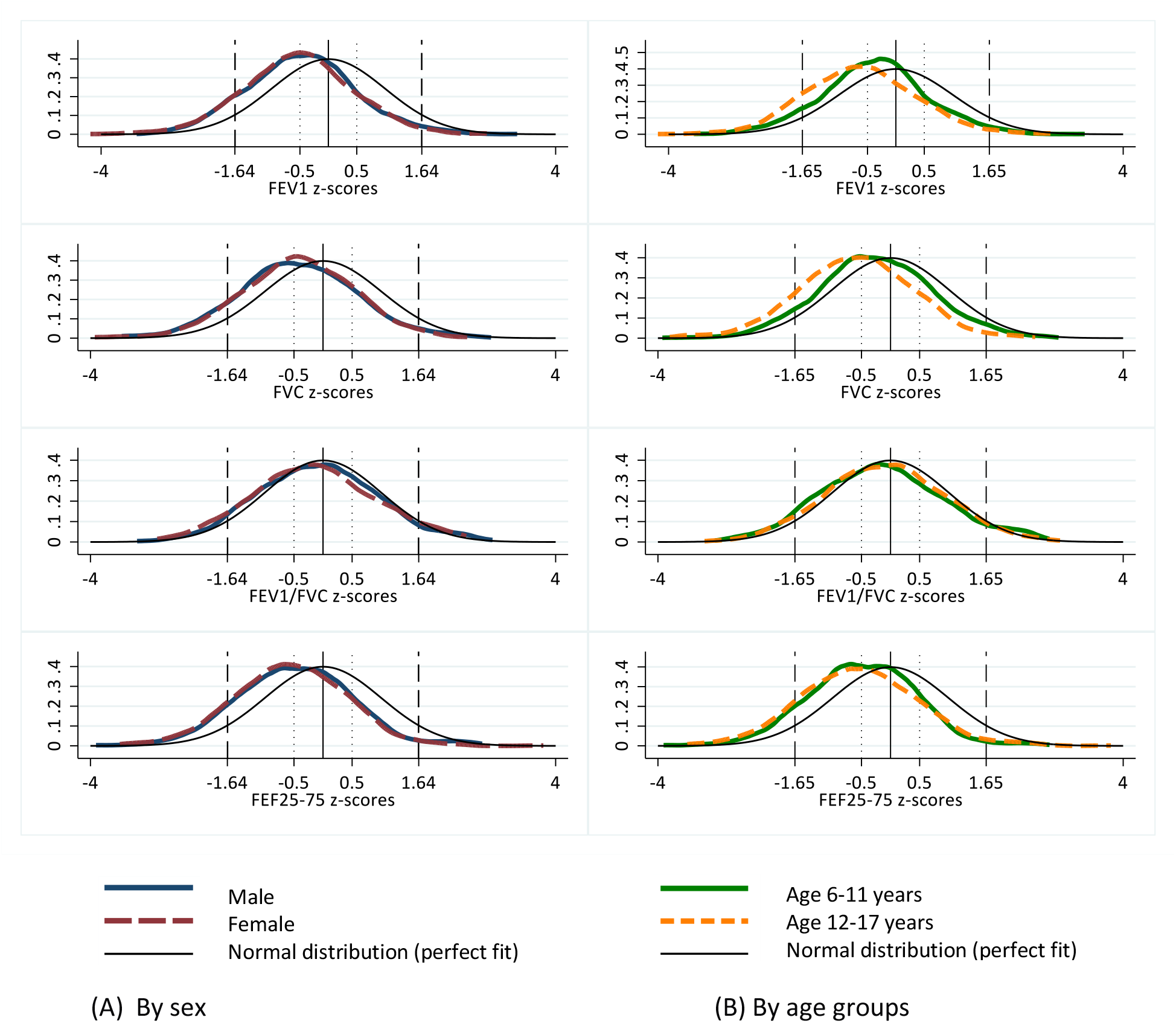
Kernel density plots showing the distribution of z-scores for spirometry parameters based on Global Lung Function Initiative (GLI) reference equations stratified by sex (A) and age groups (B) among healthy schoolchildren from the LUIS study. Dashed vertical lines mark upper and lower limits of normal (ULN, LLN) at 5% (equivalent to +1.64, -1.64 z-scores). Abbreviations: FEV_1_ = forced expiratory volume in 1 second; FVC = forced vital capacity. FEV_1_/FVC = forced expiratory volume in 1 second over forced vital capacity; FEF_25-75_ = forced expiratory flow between 25-75% of the forced vital capacity.

**Figure 3:**
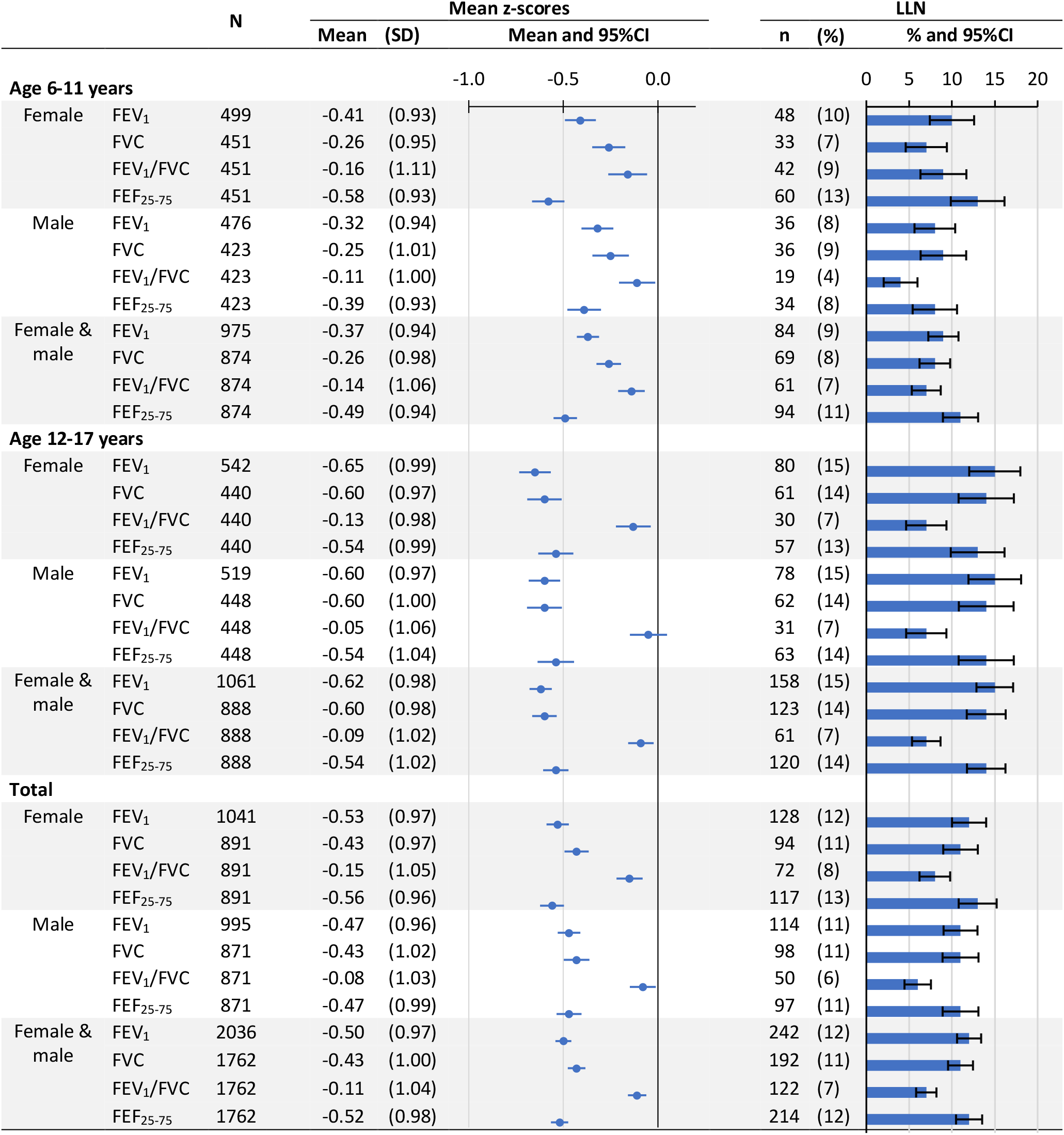
Mean spirometry GLI-based z-scores of healthy schoolchildren from the LUIS study and percentage below lower limits of normal (LLN) stratified by age and sex. LLN corresponds to z-scores <1.645. Abbreviations: GLI = Global Lung Function Initiative; FEV_1_ = forced expiratory volume in 1 second; FVC = forced vital capacity; FEV_1_/FVC = forced expiratory volume in 1 second over forced vital capacity; FEF_25-75_ = forced expiratory flow between 25-75% of forced vital capacity.

### Associations of GLI-based spirometry z-scores by age, BMI, height, and sex

FEV_1_, FVC, and FEF_25-75_ z-scores were lower for older than for younger children (Figure 4). For ages 10-11 years, FEV_1_, FVC, and FEF_25-75_ z-scores were lower for females than males (Figure 4, Table S1). FEV_1_/FVC z-scores did not vary with age (Figure 4) or height (Figure 5).

**Figure 4:**
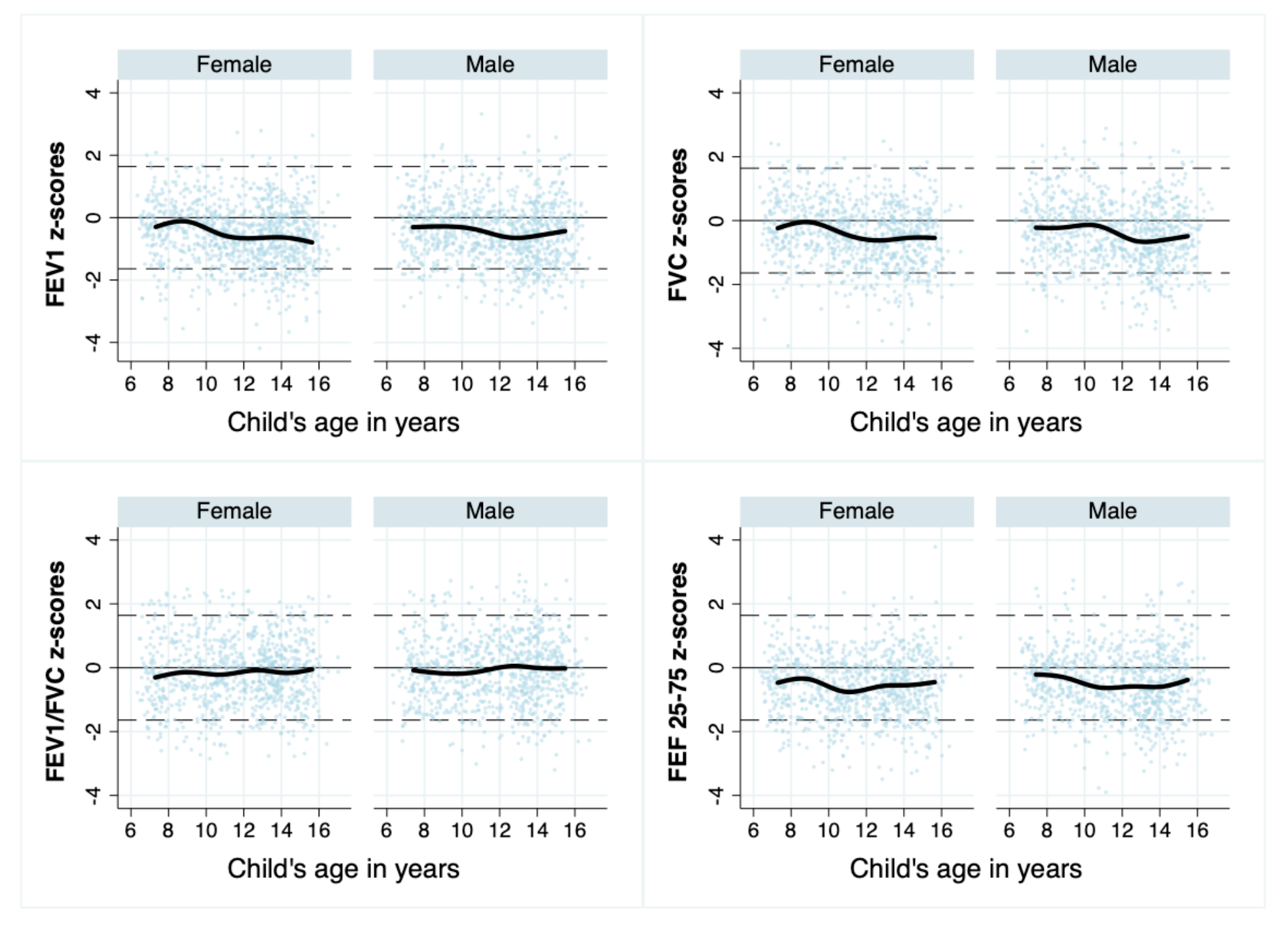
Scatter plots of GLI-based z-scores for spirometry parameters by age and by sex among healthy schoolchildren from the LUIS study. Thick black lines show median age splines for z-scores of lung funtion parameters. Dashed thin horizontal lines mark upper and lower limits of normal (ULN, LLN) at 5% (+1.64, -1.64 z-scores). Abbreviations: GLI = Global Lung Function Initiative; FEV_1_ = forced expiratory volume in 1 second; FVC = forced vital capacity; FEV_1_/FVC = forced expiratory volume in 1 second over forced vital capacity; FEF_25-75_ = forced expiratory flow between 25-75% of forced vital capacity.

**Figure 5:**
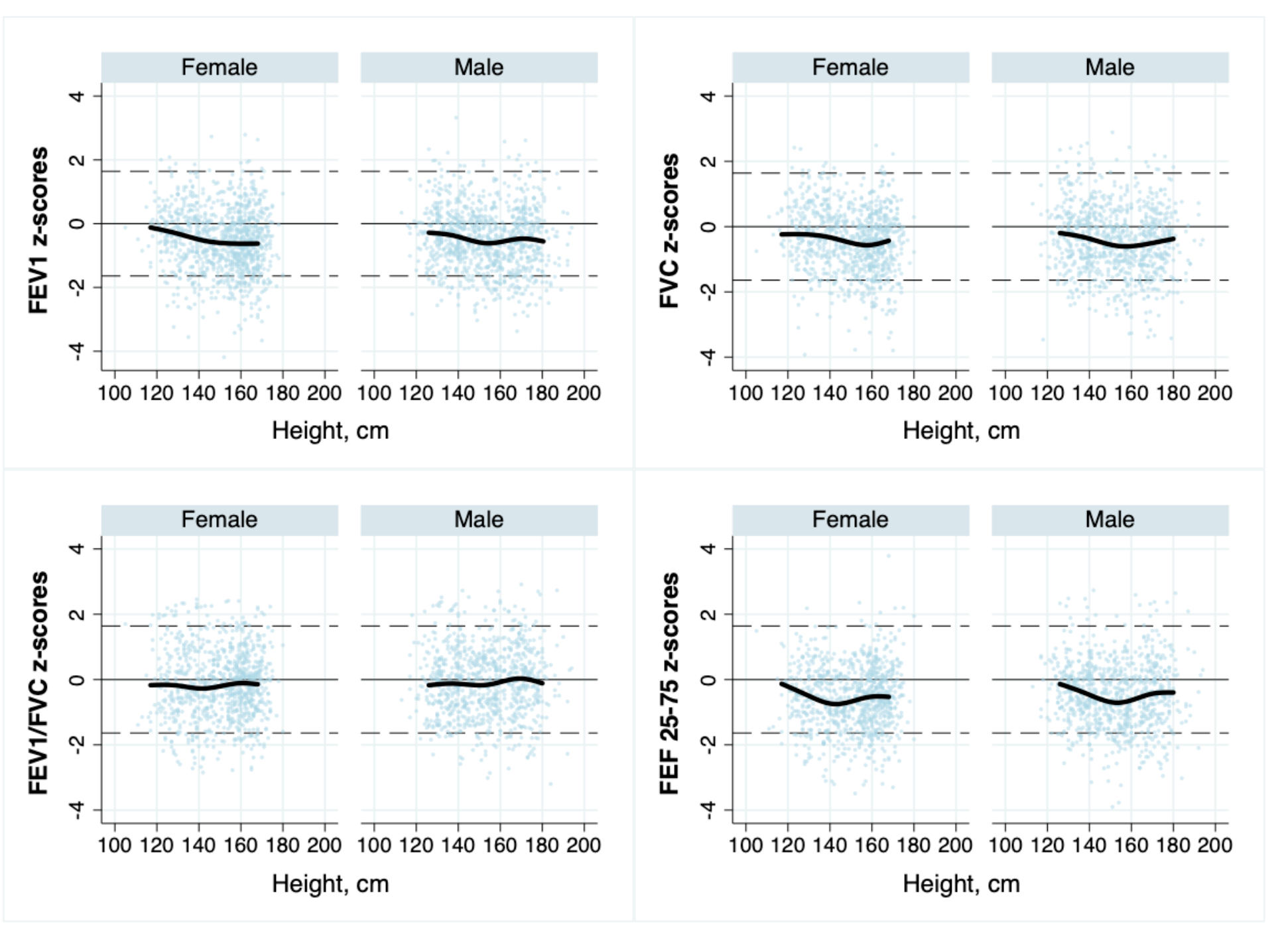
Scatter plots of GLI-based z-scores for spirometry parameters by height and by sex among healthy schoolchildren from the LUIS study. Thick black lines show median height splines for z-scores of lung funtion parameters. Dashed thin horizontal lines mark upper and lower limits of normal (ULN, LLN) at 5% (+1.64, -1.64 z-scores). Abbreviations: GLI = Global Lung Function Initiative; FEV_1_ = forced expiratory volume in 1 second; FVC = forced vital capacity; FEV_1_/FVC = forced expiratory volume in 1 second over forced vital capacity; FEF_25-75_ = forced expiratory flow between 25-75% of forced vital capacity.

There was an inverse association of FEV_1_, FVC, and FEF_25-75_ z-scores by age in multivariable linear models (Table S4) and this association with age varied by height (*p* values for interaction: <0.034, 0.019, <0.001) (Figure 6). For those aged 6-11 years, FEV_1_ and FEF_25-75_ z-scores decreased with height, while z-scores increased with height for those aged 12-17 years. FVC z-scores increased with height in all age groups, but more so for adolescents. We found no interactions with sex (data not shown).

**Figure 6:**
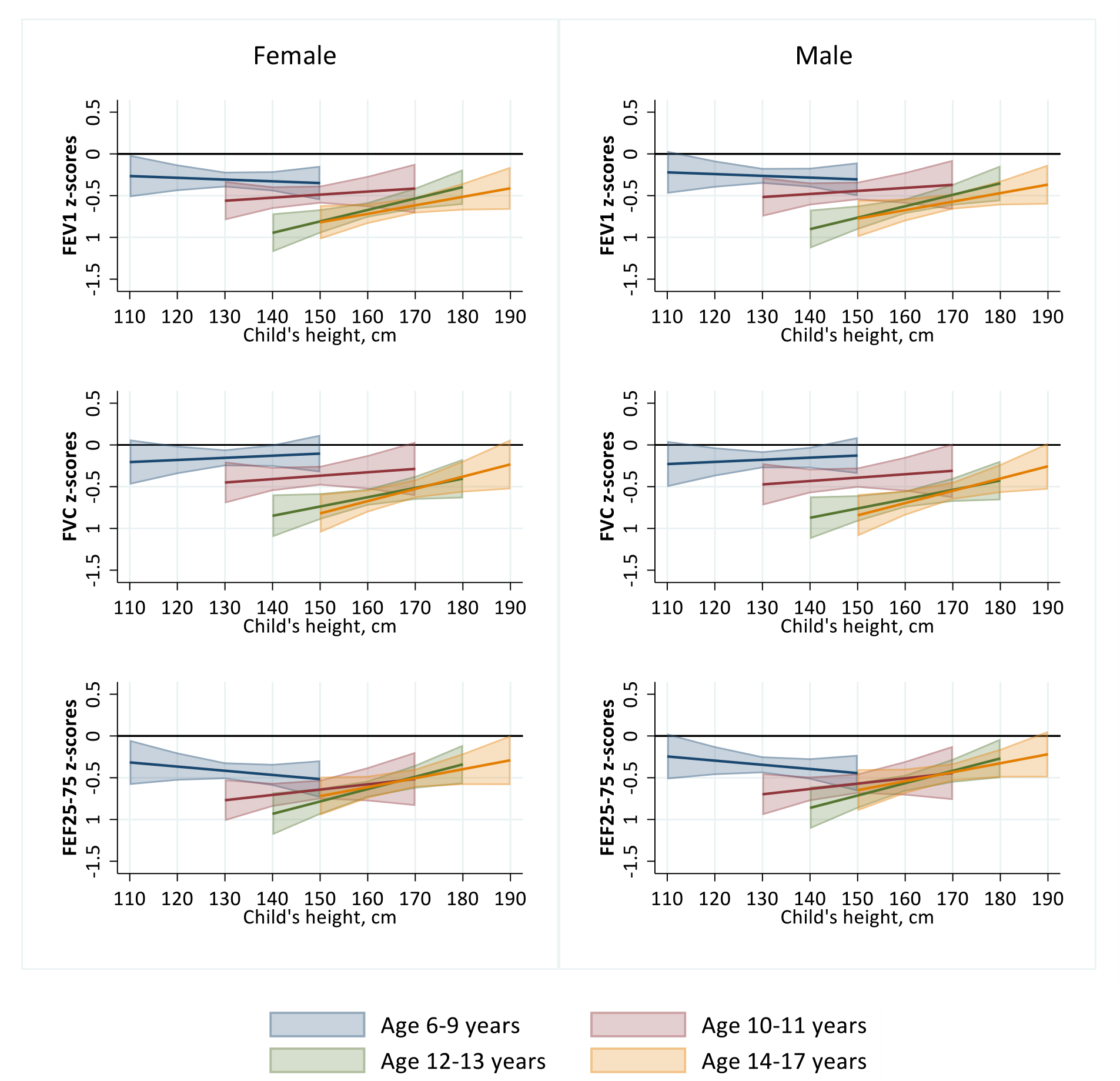
Marginal plot for predicted GLI-based z-scores for FEV1, FVC, and FEF25-75 at different heights in 4 age groups of healthy schoolchildren from the LUIS study. *P* values for interaction were produced by likelihood ratio tests comparing the models with and without the interaction term= 0.033 for FEV_1_ z-scores, 0.019 for FVC z-scores, <0.001 for FEF_25-75_ z-scores. Abbreviations: GLI = Global Lung Function Initiative; FEV_1_ = forced expiratory volume in 1 second; FVC = forced vital capacity; FEF_25-75_ = forced expiratory flow between 25-75% of forced vital capacity.

Children considered overweight or obese had higher FEV_1_, FVC, and FEF_25-75_ z-scores than children considered normal weight or underweight. Fit was poor for children classified as underweight (e.g., mean FEV_1_: -1.13; SD: 0.92) or normal weight (- 0.55; 0.96), but good for children considered overweight (−0.25; 0.92) or obese (−0.08; 0.93) (Figure 7, Table S5). FEV_1_, FVC, and FEF_25-75_ z-scores also increased with BMI in linear regression models (Table S4).

**Figure 7:**
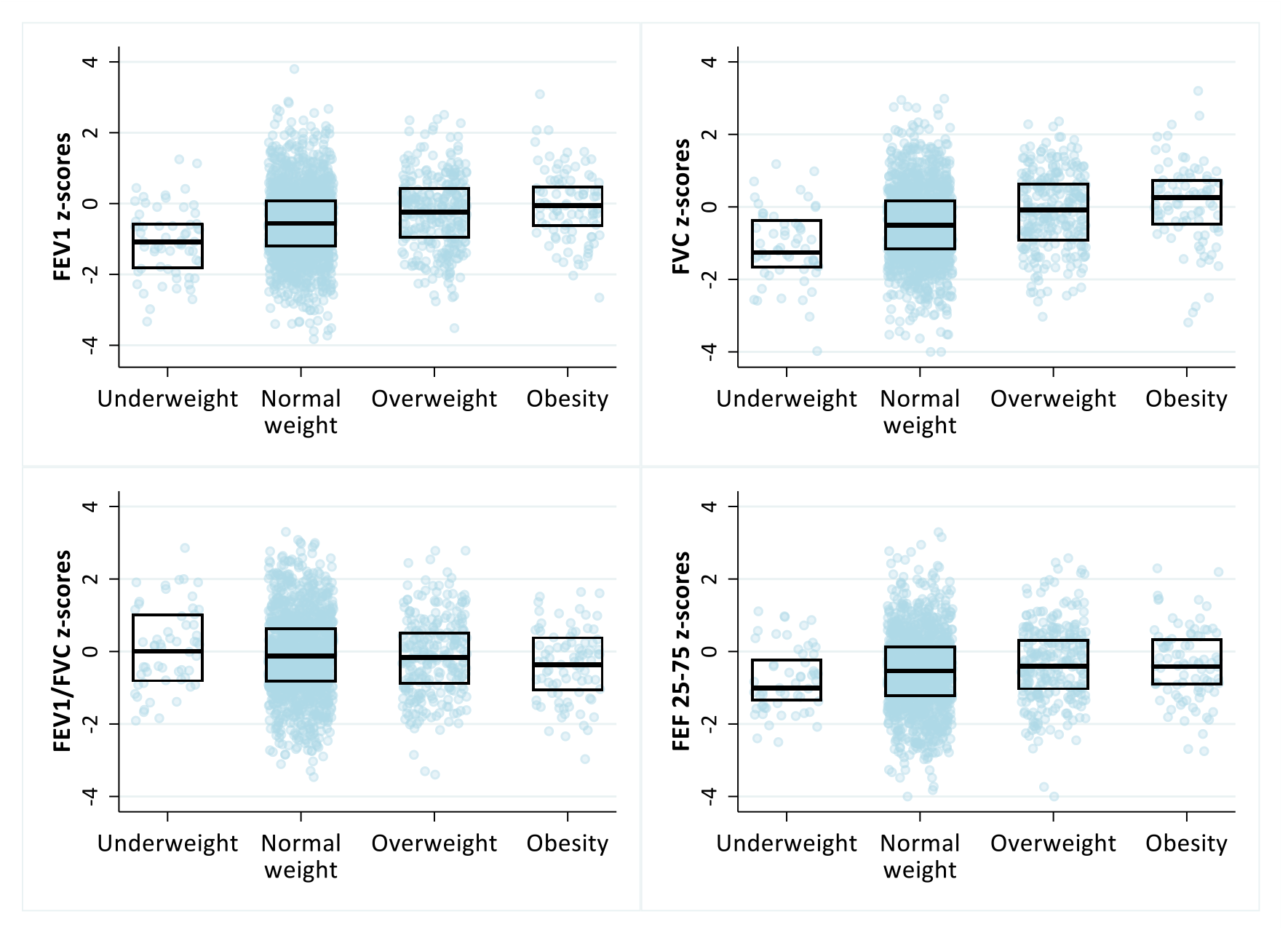
Distribution of GLI-based z-scores stratified by body mass index (BMI) categories among healthy schoolchildren from the LUIS study. *P* values for a trend in z-scores across ordered categories of BMI were <0.001 for FEV_1_, 0.014 for FVC, <0.001 for FEV_1_/FVC, and <0.001 for FEF_25-75_. The top lines of the boxes represent 75^th^ percentiles, middle lines show the median, and bottom lines the 25^th^ percentiles of GLI-based z-scores of spirometry parameters. BMI z-scores were calculated based on WHO references. BMI categories were based on WHO recommendations (underweight: <-2 z-scores; normal weight: ≥ -2, <1 z-scores; overweight: ≥ 1 z-scores; obesity: ≥2 z-scores). Abbreviations: GLI = Global Lung Function Initiative; FEV_1_ = forced expiratory volume in 1 second; FVC = forced vital capacity; FEV_1_/FVC = forced expiratory volume in 1 second over forced vital capacity; FEF_25-75_ = forced expiratory flow between 25-75% of forced vital capacity. Table S4 shows numeric results that correspond to this figure.

## Discussion

Our large population-based study of healthy schoolchildren found that FEV_1_, FVC, and FEF_25-75_ z-scores were lower than expected from GLI reference values for Swiss schoolchildren, particularly adolescents. The association of FEV_1_, FVC, and FEF_25-75_ z-scores with height differed by age. Although FEV_1_ z-scores decreased with height for younger children, they increased in older children. The fit of references for FEV_1_, FVC, and FEF_25-75_ z-scores was much better for children with higher BMI. This suggests that it is worthwhile considering whether weight or BMI data could be used to improve future GLI reference equations, resulting in a potentially better fit for samples who are slimmer or more overweight than the average.

### Strengths and limitations

Our study had several strengths. The large sample size allowed us to assess the fit of GLI-based lung function z-scores in 2 age groups of White schoolchildren each with well over 150 males and 150 females, according to GLI recommendations [1]. We performed spirometry using a state of the art set-up instead of hand-held portable spirometers and did careful quality control of spirometry flow-volume curves, which increases the reliability of our results and makes measurement bias due to poor quality unlikely [20]. We obtained information about respiratory symptoms from both parents and children which allowed us to perform sensitivity analyses to define a healthy population, adding robustness to our findings. However, our study also had some limitations. First, we cannot exclude sampling bias because not all schools took part in the study. However, participating schools were similar to the whole canton [20]. Second, we lacked information about the onset of puberty; this information would have allowed us to better explore whether pubertal stage influences fit of GLI references in adolescents. Last, although the canton of Zurich is the most populated and diverse region of Switzerland, we cannot assess if our findings apply to all Swiss schoolchildren.

### Comparison with other studies

Other studies have assessed the fit of national data with GLI reference values. The fit was good in studies from the France, Norway, Spain, and the UK [14-17]; however, fit was poorer in studies from Germany and Italy [18, 19]. Our findings are comparable to those of Hüls et al. [19]. They assessed the applicability of GLI references for White children from several German cities using a school-based study of 1943 children aged 4-19 years and a population-based birth cohort of 1042 adolescents aged 15 years. They found sufficient fit for children younger than 10 years but systematically lower mean z-scores for FEV_1_ and FVC for children older than 10 years. In the German study, mean FEV_1_ and FVC z-scores were higher for those aged 6-9 years (e.g., mean FEV_1_ z-scores = 0.01 for females and 0.02 for males), than in our study (−0.41 for females; -0.42 for males). In contrast to our study where females had lower z-scores than males (14-17 years = -0.70 for females; -0.53 for males), older males had lower FEV_1_ and FVC z-scores than older females in the 2 German cohorts (e.g., mean FEV_1_ z-scores for those aged 15-18-years = -0.24 for females; for males -0.41). In their study and in ours, fit for FEV_1_/FVC was appropriate for all age groups. Fasola et al. found that GLI references produced mean z-scores >0.5 for FEV_1_/FVC for 1243 males (mean: 0.75) and females (0.81) aged 7-16 years from southern Italy, which led to an underestimation of children with reduced FEV_1_/FVC [18]. In contrast, we found a good fit of FEV_1_/FVC z-scores. Because mean z-scores were reduced both for FEV_1_ and FVC, the ratio of these 2 remained normal.

The poor fit of GLI references for FEV_1_, FVC, and FEF_25-75_ for adolescents from our study may be related to differences in body size or timing of pubertal growth spurts that affect lung growth. Age of puberty onset, height, and timing of growth spurt vary across European countries [30, 31]. Children in northern countries generally have higher median height and later onset of puberty compared to southern countries [30, 31]. The height range in our study population was 113-194 cm for males and 105-180 cm for females aged 6-17 years, whereas the height range in the GLI reference population was 96-199 cm for males and 100-188 cm for females aged 6-18 years [32]. The mean height-for-age of our participants was higher (mean z-score: 0.56, SD: 1.01) than the WHO references [27]. This confirms that children from the canton of Zurich are on average taller than the WHO reference population [33]. Previous studies reported a slightly higher age of peak height growth in Switzerland (13.9 years for males; 12.2 years for females) than the UK (13.6 years for males; 11.7 years for females), Canada (13.4 years for males; 11.8 years for females), and the United States (13 years for males, 11 years for females) [34-36].

Another determinant of lung function in children is body composition [6]. Children considered overweight or obese have higher values for both FEV_1_ and FVC, particularly for FVC, so that the FEV_1_/FVC ratio is lower than for normal weight children [37, 38]. This aligns with our findings and may be due to asymmetric lung growth (i.e., increased dysanapsis) for children considered overweight and obese [9].

### Implications

GLI references are too high for the lung function of healthy adolescents from our study. The GLI-based LLN underestimates dynamic lung volumes and forced flows (i.e., over detects abnormal results). This misclassification affects interpretations of lung function data in research and clinical practice [39]. In research, this affects cross-sectional studies when the lung function of children with a disease is compared to normal values. It also affects longitudinal analyses when using GLI references may make lung function in older children at follow-up appear poorer than at baseline. Clinicians commonly use spirometry to diagnose respiratory disease in children. Over detection of pulmonary restriction and obstruction may induce unnecessary treatments and surveillance burdens on patients, as well as unnecessary costs on healthcare systems [39]. For example, underestimated FEV_1_ in adolescents may lead to misdiagnosis of asthma [40]. Many clinical trials use FEV_1_ to enrol or assign individuals to an intervention. Use of GLI-based FEV_1_ z-scores would classify more children below LLN and may lead to inclusion of children with lower severity. People with chronic respiratory disease are often included in national registries [41-43]. GLI-based FEV_1_ z-scores of Swiss registry participants may wrongly seem to decline from early school-age through adolescence only because of the age-related change in fit with GLI z-score, not because their disease becomes worse. This can affect international comparisons of lung health. Clinicians and researchers interpreting GLI-based FEV_1_, FVC, and FEF_25-75_ z-scores should consider the possibility of finding results below LLN in slimmer, yet otherwise healthy children. Future studies should assess ways to compensate for BMI on spirometry indices.

In conclusion, we found evidence that GLI-based spirometry reference equations fit data for White children aged 6-11 years, but not for adolescents aged 12-17 years in Switzerland. BMI is a possible driver of deviations in FEV_1_, FVC, and FEF_25-75_ z-scores and future longitudinal studies should assess the impact of timing of pubertal growth spurt on these indices. We also advise caution when interpreting borderline spirometry results in slimmer adolescents.

## Data Availability

All data produced in the present study are available upon reasonable request to the authors.

## Acknowledgements

We thank the staff from the schools, the children, and their families for taking part in the study, as well as LUIS study fieldworkers for their technical support during the study. We thank Marcel Zwahlen for his statistical advice. We thank Johanna M. Kurz. Andras Soti, Marc-Alexander Oestreich, Corin Willers (Division of Paediatric Respiratory Medicine and Allergology, Department of Paediatrics, Inselspital, Bern University Hospital, University of Bern, Switzerland), Léonie Hüsler, Eugénie Collaud, and Carmen C. M. de Jong (Institute of Social and Preventive Medicine, University of Bern, Switzerland) for their help in the assessment of the quality of the spirometry flow-volume curves. We thank Kristin Marie Bivens for her editorial contributions.

## Author’s contributions

Claudia E. Kuehni, Alexander Moeller, and Phillip Latzin conceptualised and designed the study. Alexander Moeller supervised data collection. Rebeca Mozun analysed the data and drafted the manuscript. Cristina Ardura-Garcia and Eva S.L. Pedersen supported the statistical analysis. All authors gave input for interpretation of the data. All authors critically revised and approved the manuscript.

## The LuftiBus in the school (LUIS) Study Group

Alexander Moeller, Jakob Usemann (Division of Respiratory Medicine, University Children’s Hospital Zurich and Childhood Research Centre, University of Zurich, Switzerland); Philipp Latzin, Florian Singer, and Johanna M. Kurz (Division of Paediatric Respiratory Medicine and Allergology, Department of Paediatrics, Inselspital, Bern University Hospital, University of Bern, Switzerland); Claudia E. Kuehni, Rebeca Mozun, Cristina Ardura-Garcia, Myrofora Goutaki, Eva S.L. Pedersen, and Maria Christina Mallet (Institute of Social and Preventive Medicine, University of Bern, Switzerland); Kees de Hoogh (Swiss Tropical and Public Health Institute, Basel, Switzerland).

