## Supplementary material for "Age and body mass index affect fit of spirometry GLI references in schoolchildren": Online supplement

### Online supplementary material

**Table S1:** Mean GLI-based z-scores for spirometry parameters and proportion below lower limits of normal (LLN) among healthy White Swiss schoolchildren from the LUIS study stratified by 4 age groups and by sex.

| | | Main analysis | | | |
|---|---|---|---|---|---|
| | | **N** | **Mean** | **(SD)** | **LLN n(%)** |
| **Age 6-9 y** | | | | | |
| Female | $FEV_1$ | 279 | -0.28 | (0.98) | 23 (8) |
| | FVC | 253 | -0.11 | (0.96) | 12 (5) |
| | $FEV_1$/FVC | 253 | -0.16 | (1.14) | 25 (10) |
| | $FEF_{25-75}$ | 253 | -0.46 | (0.95) | 29 (11) |
| Male | $FEV_1$ | 290 | -0.30 | (0.91) | 20 (7) |
| | FVC | 260 | -0.21 | (1.02) | 23 (9) |
| | $FEV_1$/FVC | 260 | -0.11 | (1.02) | 9 (3) |
| | $FEF_{25-75}$ | 260 | -0.33 | (0.90) | 17 (7) |
| **Age 10-11 y** | | | | | |
| Female | $FEV_1$ | 220 | -0.57 | (0.85) | 25 (11) |
| | FVC | 198 | -0.46 | (0.89) | 12(5) |
| | $FEV_1$/FVC | 198 | -0.17 | (1.08) | 17 (9) |
| | $FEF_{25-75}$ | 198 | -0.74 | (0.88) | 31 (16) |
| Male | $FEV_1$ | 186 | -0.36 | (0.98) | 16 (9) |
| | FVC | 163 | -0.31 | (1.01) | 23 (9) |
| | $FEV_1$/FVC | 163 | -0.09 | (0.98) | 10 (6) |
| | $FEF_{25-75}$ | 163 | -0.49 | (0.98) | 17 (10) |
| **Age 12-13y** | | | | | |
| Female | $FEV_1$ | 280 | -0.60 | (1.00) | 38 (14) |
| | FVC | 235 | -0.58 | (0.99) | 21 (11) |
| | $FEV_1$/FVC | 235 | -0.12 | (0.94) | 16 (6) |
| | $FEF_{25-75}$ | 235 | -0.55 | (0.94) | 28 (12) |
| Male | $FEV_1$ | 268 | -0.67 | (0.90) | 42 (16) |
| | FVC | 240 | -0.68 | (0.98) | 13 (8) |
| | $FEV_1$/FVC | 240 | -0.05 | (1.09) | 16 (7) |
| | $FEF_{25-75}$ | 240 | -0.63 | (1.00) | 34 (14) |
| **Age 14-17 y** | | | | | |
| Female | $FEV_1$ | 262 | -0.70 | (0.98) | 42 (16) |
| | FVC | 205 | -0.63 | (0.96) | 32 (16) |
| | $FEV_1$/FVC | 205 | -0.15 | (1.04) | 16 (8) |
| | $FEF_{25-75}$ | 205 | -0.53 | (1.05) | 29 (14) |
| Male | $FEV_1$ | 252 | -0.53 | (1.02) | 36 (15) |
| | FVC | 208 | -0.51 | (1.01) | 27 (13) |
| | $FEV_1$/FVC | 208 | -0.05 | (1.02) | 15 (7) |
| | $FEF_{25-75}$ | 208 | -0.45 | (1.08) | 29 (14) |

**Table S2:** Sensitivity analyses for the assessment of the fit of Global Lung Funtion Initiative (GLI) references in the LUIS study.

| Exclusion criteria | Main analysis (N=2036) | Sensitivity analysis 1 (N=2594) | Sensitivity analysis 2 (N=1150) | Sensitivity analysis 3 (N=1200) |
|---|---|---|---|---|
| **Definition of healthy** | | | | |
| Questionnaire missing | | | | |
| Parent questionnaire | x | | x | x |
| Child questionnaire | x | | x | x |
| Wheeze (past 12m) | | | | |
| Parent reported | x | x | x | x |
| Child reported | | | x | |
| Asthma diagnosis (ever) | | | | |
| Parent reported | x | | x | x |
| Child reported | | | x | |
| Inhaled corticosteroid use (past 12m)* | x | x | x | x |
| Chronic cough (past 12m)* | | | x | |
| Active smoking** | | | | |
| Everyday | x | x | x | x |
| >=once/week | x | | x | x |
| <once/week | | | x | |
| Sick on the day of measurement** | | | | |
| Strong cough or cold | x | x | x | x |
| Mild cough or cold | | | x | |

*Information only available from the parent questionnaire. **Information only available from the child questionnaire. Sensitivity analyses: (1) more inclusive sample selection approach as in the main analysis; (2) more restrictive sample selection approach as in the main analysis; (3) sample selection approach as in the main analysis, but additionally excluding children with migration background. "x" indicates the exclusion criteria for each analysis.

**Table S3:** Mean GLI-based z-scores for spirometry parameters and proportion below lower limits of normal (LLN) among healthy Swiss schoolchildren from the LUIS study by age and by sex for the main and sensitivity analyses.

| | | Main analysis | | | | Sensitivity analysis 1 | | | | Sensitivity analysis 2 | | | | Sensitivity analysis 3 | | | |
|---|---|---|---|---|---|---|---|---|---|---|---|---|---|---|---|---|---|
| | | N | Mean | (SD) | LLN n (%) | N | Mean | (SD) | LLN n (%) | N | Mean | (SD) | LLN n (%) | N | Mean | (SD) | LLN n (%) |
| **Age 6-11 years** | | | | | | | | | | | | | | | | | |
| F | $FEV_1$ | 499 | -0.41 | (0.93) | 48 (10) | 534 | -0.42 | (0.95) | 55 (10) | 248 | -0.44 | (0.93) | 25 (10) | 312 | -0.37 | (0.90) | 26 (8) |
| | FVC | 451 | -0.26 | (0.95) | 33 (7) | 479 | -0.25 | (0.96) | 36 (8) | 219 | -0.29 | (0.99) | 20 (9) | 288 | -0.23 | (0.90) | 17 (6) |
| | $FEV_1$/FVC | 451 | -0.16 | (1.11) | 42 (9) | 479 | -0.20 | (1.11) | 46 (10) | 219 | -0.14 | (1.11) | 20 (9) | 288 | -0.18 | (1.09) | 27 (9) |
| | $FEF_{25-75}$ | 451 | -0.58 | (0.93) | 60 (13) | 479 | -0.61 | (0.93) | 67 (14) | 219 | -0.61 | (0.89) | 26 (12) | 288 | -0.60 | (0.90) | 35 (12) |
| M | $FEV_1$ | 476 | -0.32 | (0.94) | 36 (8) | 524 | -0.34 | (0.95) | 42 (8) | 246 | -0.25 | (0.94) | 15 (6) | 292 | -0.35 | (0.92) | 21 (7) |
| | FVC | 423 | -0.25 | (1.01) | 36 (9) | 465 | -0.24 | (1.01) | 39 (8) | 219 | -0.16 | (1.03) | 15 (7) | 262 | -0.25 | (0.97) | 21 (8) |
| | $FEV_1$/FVC | 423 | -0.11 | (1.00) | 19 (4) | 465 | -0.14 | (1.00) | 24 (5) | 219 | -0.09 | (1.03) | 10 (5) | 262 | -0.16 | (1.02) | 12 (5) |
| | $FEF_{25-75}$ | 423 | -0.39 | (0.93) | 34 (8) | 465 | -0.43 | (0.94) | 40 (9) | 219 | -0.34 | (0.95) | 14 (6) | 262 | -0.45 | (0.90) | 17 (6) |
| **Age 12-17 years** | | | | | | | | | | | | | | | | | |
| F | $FEV_1$ | 542 | -0.65 | (0.99) | 80 (15) | 752 | -0.66 | (0.99) | 109 (14) | 335 | -0.59 | (0.98) | 45 (13) | 301 | -0.65 | (1.01) | 43 (14) |
| | FVC | 440 | -0.60 | (0.97) | 61 (14) | 596 | -0.59 | (0.98) | 80 (13) | 274 | -0.55 | (0.97) | 36 (13) | 246 | -0.63 | (0.98) | 35 (14) |
| | $FEV_1$/FVC | 440 | -0.13 | (0.98) | 30 (7) | 596 | -0.19 | (1.00) | 49 (8) | 274 | -0.11 | (0.98) | 17 (6) | 246 | -0.13 | (0.98) | 17 (7) |
| | $FEF_{25-75}$ | 440 | -0.54 | (0.99) | 57 (13) | 596 | -0.57 | (1.01) | 83 (14) | 274 | -0.51 | (0.98) | 33 (12) | 246 | -0.59 | (1.02) | 33 (13) |
| M | $FEV_1$ | 519 | -0.60 | (0.97) | 78 (15) | 784 | -0.57 | (0.97) | 111 (14) | 321 | -0.64 | (0.94) | 50 (16) | 295 | -0.62 | (0.94) | 47 (16) |
| | FVC | 448 | -0.60 | (1.00) | 62 (14) | 653 | -0.56 | (0.99) | 88 (13) | 278 | -0.67 | (0.97) | 44 (16) | 257 | -0.57 | (1.00) | 32 (12) |
| | $FEV_1$/FVC | 448 | -0.05 | (1.06) | 31 (7) | 653 | -0.10 | (1.07) | 50 (8) | 278 | 0.00 | (1.03) | 15 (5) | 257 | -0.09 | (1.09) | 22 (9) |
| | $FEF_{25-75}$ | 448 | -0.54 | (1.04) | 63 (14) | 653 | -0.56 | (1.06) | 100 (15) | 278 | -0.52 | (1.03) | 38 (14) | 257 | -0.60 | (1.00) | 37 (14) |

LLN corresponds to z-scores <1.645. Sensitivity analyses: (1) more inclusive sample selection approach as in the main analysis; (2) more restrictive sample selection approach as in the main analysis; (3) sample selection approach as in the main analysis, but additionally excluding children with migration backgrounds. See table S1 for more detailed information on the sensitivity analyses. Abbreviations: GLI = Global Lung Function Initiative; F = female; M = male; FEV1 = forced expiratory volume in 1 second; FVC = forced vital capacity; FEV1/FVC = forced expiratory volume in 1 second over forced vital capacity; $FEF_{25-75}$ = forced expiratory flow between 25-75% of forced vital capacity.

**Table S4**: Association of GLI-based z-scores of spirometry parameters with age, body mass index (BMI), height, and sex for healthy Swiss schoolchildren from the LUIS study using linear regression models.

(a) Univariable

| | $FEV_1$ z-scores | | FVC z-scores | | $FEV_1$/FVC z-scores | | $FEF_{25-75}$ z-scores | |
|---|---|---|---|---|---|---|---|---|
| | Coeff. | [95% CI] | Coeff. | [95% CI] | Coeff. | [95% CI] | Coeff. | [95% CI] |
| Age, per year increase | -0.05 | [-0.06,-0.03] | -0.06 | [-0.08,-0.05] | 0.01 | [-0.01,0.03] | -0.01 | [-0.03,0.00] |
| Height, per 10cm increase | -0.06 | [-0.08,-0.06] | -0.08 | [-0.11,-0.05] | 0.01 | [-0.02,0.04] | 0.01 | [-0.04,0.02] |
| Sex, male | 0.07 | [-0.02,0.15] | 0.00 | [-0.09,0.09] | 0.07 | [-0.03,0.17] | 0.09 | [0.00,0.18] |
| BMI, per 1 z-score increase | 0.22 | [0.18,0.25] | 0.25 | [0.21,0.29] | -0.07 | [-0.11,-0.03] | 0.11 | [0.07,0.15] |
| N | 2036 | | 1762 | | 1762 | | 1762 | |

(b) Multivariable model 1

| | $FEV_1$ z-scores | | FVC z-scores | | $FEV_1$/FVC z-scores | | $FEF_{25-75}$ z-scores | |
|---|---|---|---|---|---|---|---|---|
| | Coeff. | [95% CI] | Coeff. | [95% CI] | Coeff. | [95% CI] | Coeff. | [95% CI] |
| Age, per year increase | -0.26 | [-0.42,-0.09] | -0.32 | [-0.50,-0.13] | 0.02 | [-0.03,0.06] | -0.37 | [-0.56,-0.19] |
| Height, per 10cm increase | -0.09 | [-0.24,0.06] | -0.11 | [-0.27,0.05] | -0.02 | [-0.08,0.05] | -0.22 | [-0.38,-0.06] |
| Age * Height interaction | 0.01 | [0.00,0.02] | 0.01 | [0.00,0.03] | | - | 0.02 | [0.01,0.03] |
| Sex, male | 0.05 | [-0.04,0.13] | -0.01 | [-0.11,0.08] | 0.07 | [-0.02,0.17] | 0.07 | [-0.02,0.16] |
| N | 2036 | | 1762 | | 1762 | | 1762 | |
| Adjusted R-squared | 0.020 | | 0.031 | | 0.000 | | 0.010 | |

(c) Multivariable model 2

| | $FEV_1$ z-scores | | FVC z-scores | | $FEV_1$/FVC z-scores | | $FEF_{25-75}$ z-scores | |
|---|---|---|---|---|---|---|---|---|
| | Coeff. | [95% CI] | Coeff. | [95% CI] | Coeff. | [95% CI] | Coeff. | [95% CI] |
| Age, per year increase | -0.22 | [-0.38,-0.06] | -0.26 | [-0.44,-0.08] | 0.01 | [-0.03,0.05] | -0.35 | [-0.53,-0.17] |
| Sex, male | 0.04 | [-0.05,0.12] | -0.02 | [-0.11,0.07] | 0.08 | [-0.02,0.17] | 0.07 | [-0.02,0.16] |
| Height, per 10cm increase | -0.14 | [-0.29,-0.00] | -0.18 | [-0.33,-0.02] | 0.01 | [-0.06,0.08] | -0.24 | [-0.40,-0.09] |
| Age * Height interaction | 0.01 | [0.00,0.02] | 0.01 | [0.00,0.03] | | - | 0.02 | [0.01,0.03] |
| BMI, per 1 z-score increase | 0.23 | [0.19,0.27] | 0.27 | [0.23,0.31] | -0.08 | [-0.12,-0.03] | 0.11 | [0.07,0.15] |
| N | 2036 | | 1762 | | 1762 | | 1762 | |
| Adjusted R-squared | 0.085 | | 0.120 | | 0.006 | | 0.025 | |

Multivariable linear regressions were adjusted for all variables in the table and included an interaction term for the outcomes $FEV_1$, FVC, and $FEF_{25-75}$. *P* values for interaction produced by likelihood ratio tests between the models with and without the interaction term in model (b) = 0.033 for $FEV_1$, 0.019 for FVC, 0.879 for $FEV_1$/FVC, and <0.001 for $FEF_{25-75}$; and in model (c) = 0.040 for $FEV_1$, 0.026 for FVC, 0.831 for $FEV_1$/FVC, and <0.001 for $FEF_{25-75}$. BMI z-scores were calculated based on WHO references. Abbreviations: CI = confidence intervals; GLI = Global Lung Function Initiative; $FEV_1$ = forced expiratory volume in 1 second; FVC = forced vital capacity; $FEV_1$/FVC = forced expiratory volume in 1 second over forced vital capacity; $FEF_{25-75}$ = forced expiratory flow between 25-75% of forced vital capacity.

**Table S5:** Mean GLI-based z-scores for spirometry parameters stratified by body mass index (BMI) groups among healthy Swiss schoolchildren from the LUIS study.

(a) BMI categories

| | $FEV_1$ z-scores | | | FVC z-scores | | | $FEV_1$/FVC z-scores | | $FEF_{25-75}$ z-scores | |
|---|---|---|---|---|---|---|---|---|---|---|
| | N | mean | (SD) | N | mean | (SD) | mean | (SD) | mean | (SD) |
| Underweight | 61 | -1.13 | (0.92) | 56 | -1.14 | (1.01) | 0.06 | (1.01) | -0.82 | (0.84) |
| Normal weight | 1573 | -0.55 | (0.96) | 1346 | -0.50 | (0.98) | -0.09 | (1.06) | -0.55 | (0.98) |
| Overweight | 310 | -0.25 | (0.92) | 274 | -0.13 | (0.92) | -0.19 | (1.01) | -0.38 | (1.00) |
| Obese | 92 | -0.08 | (0.93) | 86 | 0.12 | (0.99) | -0.31 | (0.88) | -0.30 | (0.91) |
| p trend | | <0.001 | | | 0.014 | | <0.001 | | <0.001 | |

BMI z-scores were calculated based on WHO references and categories were based on WHO recommendations (underweight: <-2 z-scores; normal weight: ≥ -2, <1 z-scores; overweight: ≥ 1 z-scores; obese: ≥2 z-scores).

(b) BMI quartiles

| | $FEV_1$ z-scores | | | FVC z-scores | | | $FEV_1$/FVC z-scores | | $FEF_{25-75}$ z-scores | |
|---|---|---|---|---|---|---|---|---|---|---|
| | N | mean | (SD) | N | mean | (SD) | mean | (SD) | mean | (SD) |
| 1st quartile | 509 | -0.83 | (0.92) | 431 | -0.84 | (0.98) | 0.02 | (1.10) | -0.66 | (0.96) |
| 2nd quartile | 513 | -0.54 | (0.88) | 431 | -0.48 | (0.91) | -0.14 | (1.02) | -0.59 | (0.92) |
| 3rd quartile | 507 | -0.40 | (1.00) | 449 | -0.32 | (0.98) | -0.13 | (1.03) | -0.46 | (1.00) |
| 4th quartile | 507 | -0.22 | (0.97) | 451 | -0.10 | (0.97) | -0.21 | (1.01) | -0.36 | (1.01) |
| p trend | | <0.001 | | | <0.001 | | 0.003 | | <0.001 | |

Quartiles ordered from lowest ($1^{st}$ quartile) to highest ($4^{th}$ quartile) BMI z-scores.

Abbreviations: GLI = Global Lung Function Initiative; $FEV_1$ = forced expiratory volume in 1 second. FVC = forced vital capacity; $FEV_1$/FVC = forced expiratory volume in 1 second over forced vital capacity; $FEF_{25-75}$ = forced expiratory flow between 25-75% of forced vital capacity; *p* trend = *p* value for a trend in z-scores of spirometry parameters across ordered quartiles of BMI.

**Table S6:** Results of different European studies that assessed the fit of Global Lung Function Initiave (GLI) references for healthy children.

| | Hüls et al. [1] | Fasola et al. [2] | Bonner et al. [3] | Nevè et al. [4] | Martín de Vicente et al. [5] | Langhammer et al. [6] |
|---|---|---|---|---|---|---|
| **Country** | Germany | Italy | UK | France | Spain | Norway |
| **Region** | LUNOKID: Wesel, Düsseldorf, Hannover. / GINIplus: Munich and Wesel | Sicily | London | Nord-Pas de Calais region of northern France | Barcelona | |
| **Years of study conduct** | 2008-2009 / 1995-1998 | 2004-2011 | 2010-2012 | 2006-2007 | - | 1995-2008 |
| **Exclusion criteria (definition of healthy)** | respiratory disease; diagnosed asthma; spastic bronchitis | wheeze ever, current wheeze, dry night cough apart from colds, doctor-diagnosed asthma, rhinitis apart from colds | current asthma; obstruction in spirometry; sickle cell disease | respiratory disease | repeated episodes of bronchitis, chronic lung disease; systemic disease with pulmonary involvement; heart, neuromuscular, or bone disease | self-reported respiratory disease, respiratory symptoms, |
| | active smoking | - | - | smoking | - | smoking history |
| | infection on the day of the measurement; lower respiratory tract infection past 6 weeks. | respiratory tract infections in the past 12 months | symptomatic on the test day | respiratory infection month before the tests | - | - |
| | * | birth weight <2000g | preterm | - | preterm | - |
| | - | BMI z-score > 2 or <-2; Both parents non-European | - | - | - | - |
| **Age range, years** | LUNOKID: 4-19 / GINIplus: 15 | 7-16 | 5-11 | 3-15 | 3-6 | 12-19 |
| **N** | 3205 / 1628 | 1243 | 1088 (359 white) | 442 | 380 | 8725 |
| **Mean z-scores $FEV_1$** | Boys: -0.26; Girls: -0.17 / Boys: -0.62; Girls: -0.55 | Boys: 0.15; Girls: 0.07 | Boys and girls: 0.03 | Boys: 0.31, Girls: 0.30 | Boys: 0.11; Girls: -0.06 | Boys: 0.02; Girls: -0.07 |
| **Mean z-scores FVC** | Boys: -0.16; Girls: -0.04 / Boys: -0.65; Girls: -0.61 | Boys: -0.17; Girls: -0.42. | Boys and girls: 0.17 | Boys: 0.19; Girls: 0.18 | Boys: -0.48; Girls: -0.42 | Boys: -0.01; Girls: -0.03 |

*Sensitivity analyses excluding children with passive smoking exposure, eczema, hay fever, and preterm birth.

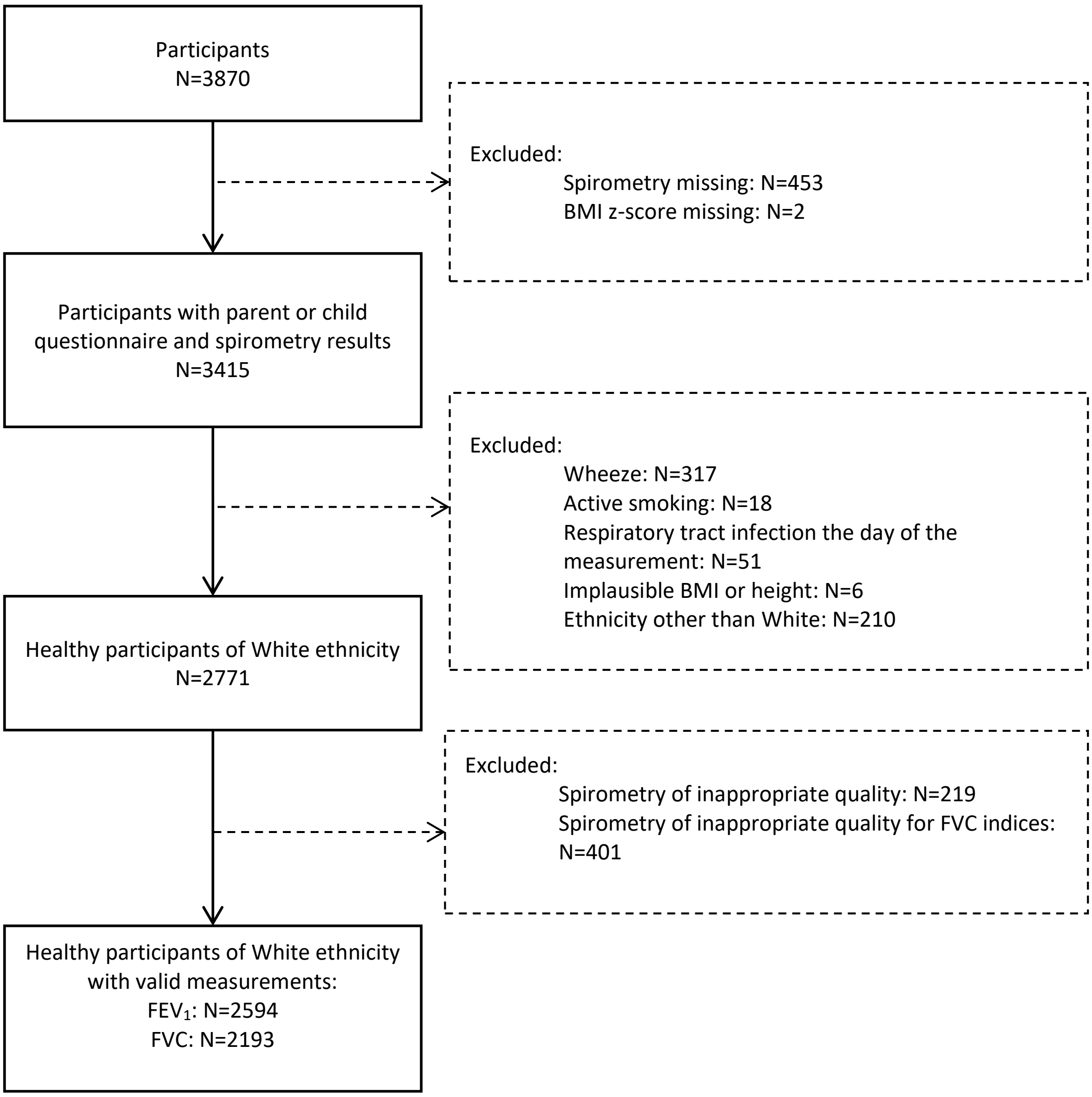

**Figure S1:** Flowchart showing the selection of the study population for the main analysis and for sensitivity analysis 1.

Wheeze: parent reported wheeze in the past 12 months or use of inhaled corticosteroids in the past 12 months. Active smoking: child reported daily smoking of cigarettes, shishas, or e-cigarettes. Sick the day of the measurement: child reported strong cough or cold the day of the measurement.

Spirometry of inappropriate quality: We additionally excluded results for FVC related parameters when we identified signs of early termination of expiration or cough or glottis closure after the first second of expiration in flow-volume curves.

Abbreviations: $FEV_1$ = forced expiratory volume in 1 second; FVC = forced vital capacity; BMI = body mass index.

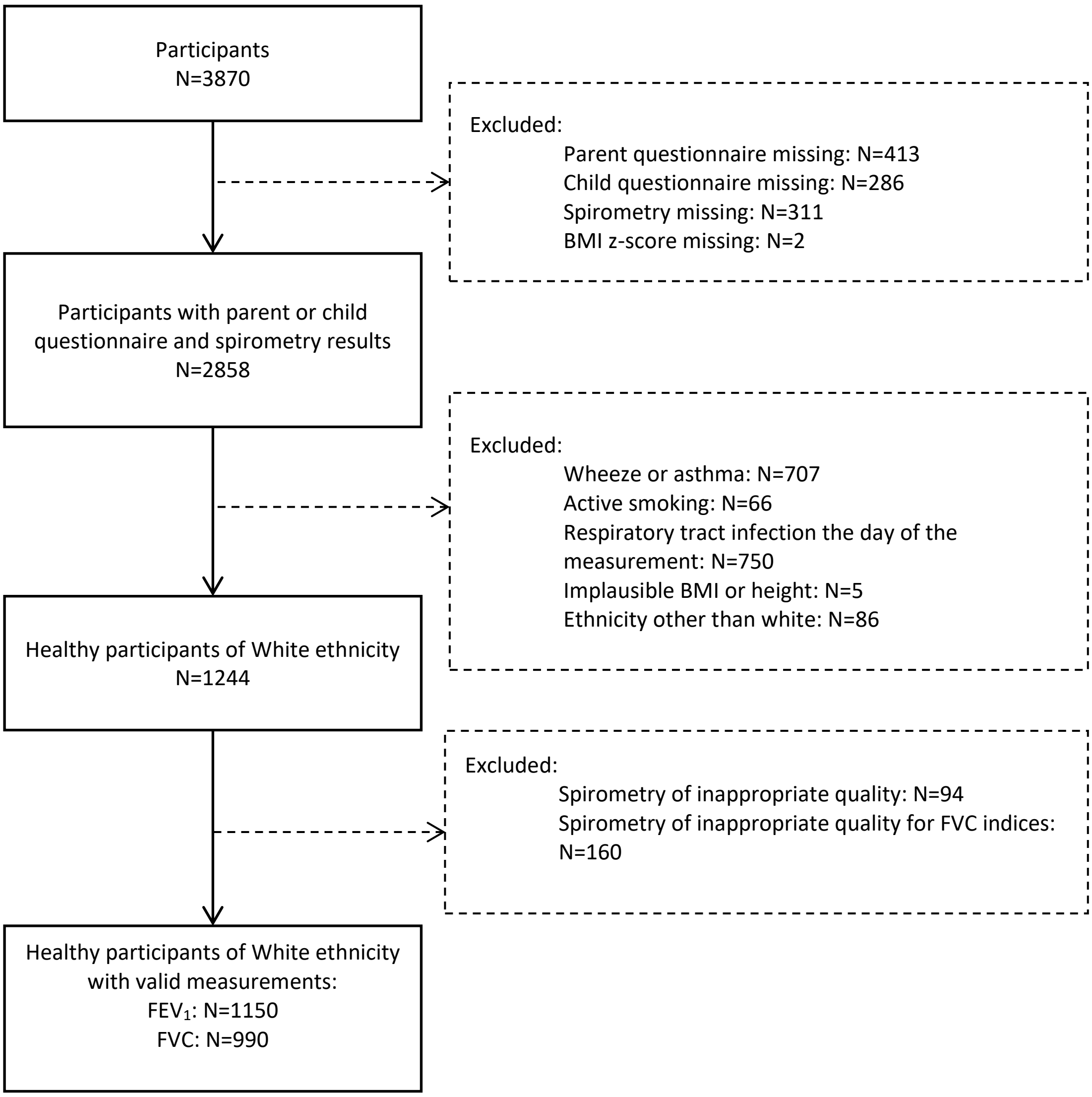

**Figure S2:** Flowchart showing the selection of the study population for the main analysis and for sensitivity analysis 2.

Wheeze or asthma: parent or child reported wheeze in the past 12 months, use of inhaled corticosteroids in the past 12 months, cough lasting longer than 3 weeks in the past 12 months, or asthma diagnosed by a physician ever in life. Active smoking: child reported smoking cigarettes, shishas, or e-cigarettes less than once per week. Sick the day of the measurement: child reported mild or strong cough or cold the day of the measurement.

Spirometry of inappropriate quality: We additionally excluded results for FVC related parameters when we identified signs of early termination of expiration or cough or glottis closure after the first second of expiration in flow-volume curves.

Abbreviations: $FEV_1$ = forced expiratory volume in 1 second. FVC = forced vital capacity. BMI = body mass index.
